# “Return to University Campuses Associated with 9% Increase in New COVID-19 Case Rate”

**DOI:** 10.1101/2020.10.13.20212183

**Authors:** Grace Bagwell Adams, Jerry Shannon, Sarah Shannon

## Abstract

The vast majority of colleges and universities across the United States are bringing students back for in-person instruction in the midst of the COVID-19 pandemic, despite the absence of an effective vaccine or other anti-viral therapeutic treatment. Using data from the New York Times and the American Community Survey, we assess the effect of this return to campus on viral case growth in counties with a significant college student population (“college counties”) relative to non-college counties. We find a significant surge in new cases in a 21-day time frame in college counties, a finding consistent across U.S. Census divisions. These results suggest the need for institutions of higher education and the communities where these institutions reside work together quickly and effectively to mitigate viral transmission and to prevent overwhelming local healthcare infrastructure in college counties.

## Introduction

In March of 2020, institutions of higher education across the United States pivoted abruptly to remote teaching in response to the COVID-19 pandemic^1^. These institutions complied with the shelter-in-place policies and public health guidance that sought to “flatten the curve,^”^ and slow the speed of virus transmission.^2^

Throughout the summer that followed, colleges and universities nationwide began to formulate plans to reopen in the fall, or to remain closed to in-person student life and instruction. While the approaches taken by these postsecondary institutions to promote public health and safety have varied, the commonality shared across schools with any in-person instruction is the significant influx of students to the communities where these institutions are located.

According to the most recent data tracked by Davidson College’s College Crisis Initiative, only 191 of 2,958 colleges and universities are fully online for fall semester 2020.^3^ This means that the vast majority of U.S. institutions of higher education are bringing students back to campus in some way, and that this fall’s return to campus is occurring without a vaccine to prevent or effective therapeutic to treat the Sars-CoV-2 virus.

This return to campus in the midst of an ongoing global pandemic brings together a disease that spreads rapidly through respiratory droplets and a population that is prone to spending substantial time in large groups, especially at bars and parties, which are high risk scenarios for COVID-19 transmission.^4,5^ Indeed, despite the best laid plans for a “safe” return to campus, already some institutions of higher education are ceasing or substantially decreasing in-person activities and pivoting back to remote instruction. Others soldier on, despite alarming increases in cases among the study body and surrounding community.

Our goal in this data analysis is to describe the initial return to campus rise in COVID-19 cases in college counties across the United States during late August and early September 2020. We present a novel measure of the “return to campus COVID19 new case rate” and provide descriptive evidence of how and where outbreaks are emerging in college counties. We estimate that counties where 10 percent or more of the population is enrolled in college experience a 9 percent increase in the new COVID19 case rate during the return to campus period as compared to other counties. This significant rise in COVID19 case rates associated with the return of college students to college towns has substantial implications for public health, especially in rural communities with already strained healthcare infrastructure and worrisome health disparities.

### Study Data & Methods

Our data come from two publicly available sources. Census divisions, the percent of the population aged 18 to 24, and total county population data come from the U.S. Census Bureau.^6^ COVID19 case counts and deaths come from the New York Times COVID19 database.^7^ COVID-19 case data are available at the county-day level, which is the unit of analysis. The observation time frame extends retrospectively, twelve weeks from the most recent case reporting available at the time of our writing, from June 14 through September 6, 2020. The county day sample size is 3,118 records.

The main outcome of interest is what we term the *new case rate*, where the number of new cases in the last three weeks prior to September 6 (August 16) was divided by the cases in the twelve weeks prior to this date (June 14). To produce a comparable scale with the main variable of interest (percentage college students), this ratio is multiplied by 100. If cases were distributed evenly, we would expect a rate of 25%, as three weeks is one-quarter of the twelve-week study period. A higher number would thus indicate a surge of new cases, while a lower one would indicate a tapering off of cases.

The independent variable of interest is a dichotomous indicator of whether the county is defined as a “college county.” To operationalize this hypothesis, we use data from the 2014-18 American Community Survey showing the number of individuals ages 18 to 24 currently enrolled in public college or graduate school. We rely on the New York Times decision rule of assigning a value of “1” to a county if at least 10% of the county’s total population is comprised of college students ages 18-24 (n=135); counties that do not meet this criterion are assigned a “0” value (n=2,983). In addition to the new case rate and the college county indicator, we use the geographic areas defined as the nine U.S. Census divisions to control for regional variation. We also control for total county population.

We estimated descriptive statistics for all variables and include those in Exhibit 2. We then estimated a regression model using the return to campus new case rate as the outcome of interest. As a sensitivity analysis, we also estimated an alternative outcome using a rate that is the return to campus new cases (the most recent 21 days) divided by total cases since the beginning of the pandemic. We also created a linear model for just college counties including the percentage of college students along with census division and total population. All estimates were conducted using R v.3.6.0.

**Exhibit 1:**
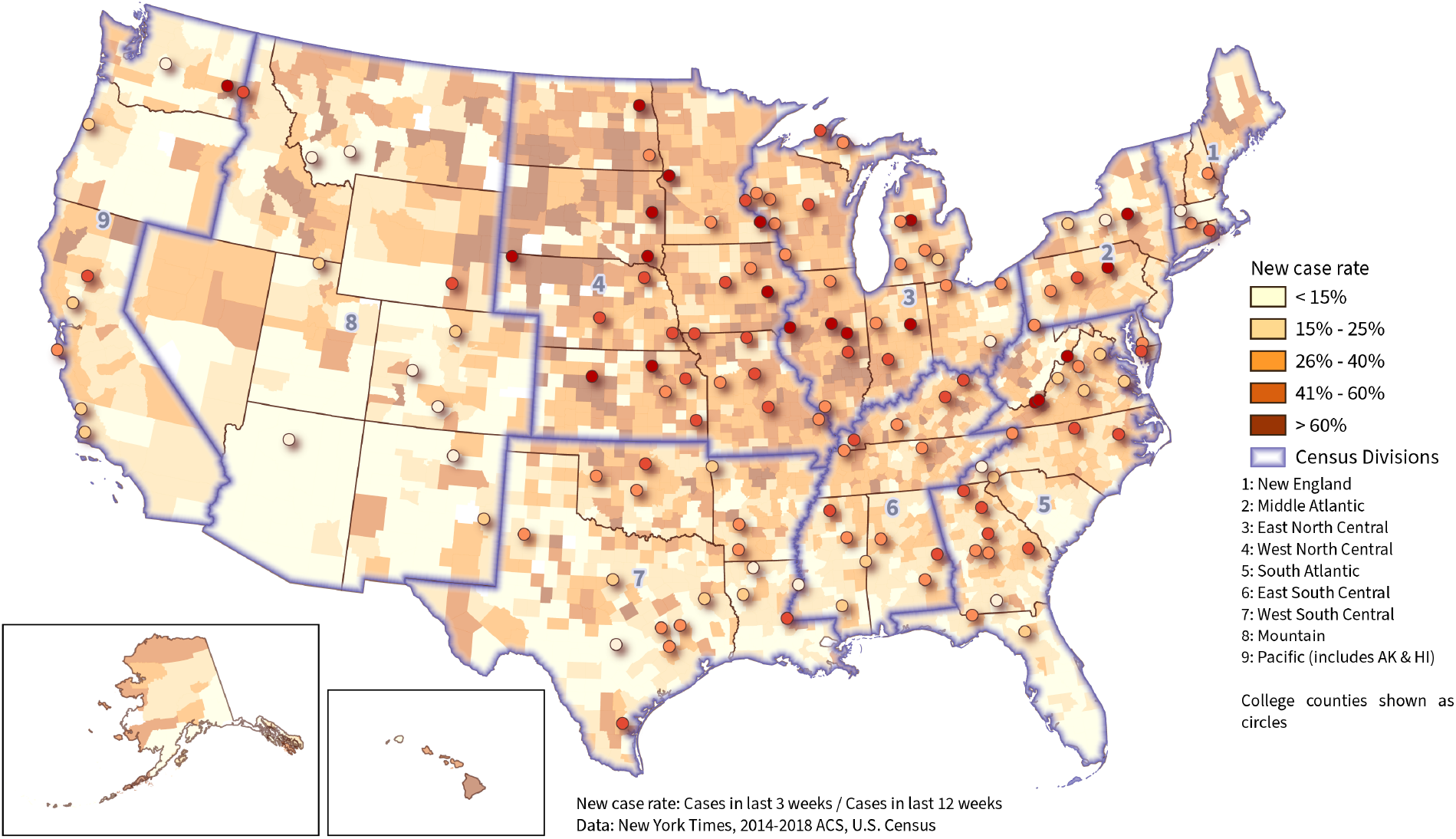
U.S. Map of Census Divisions & COVID19 Case Growth.

**Exhibit 2:**
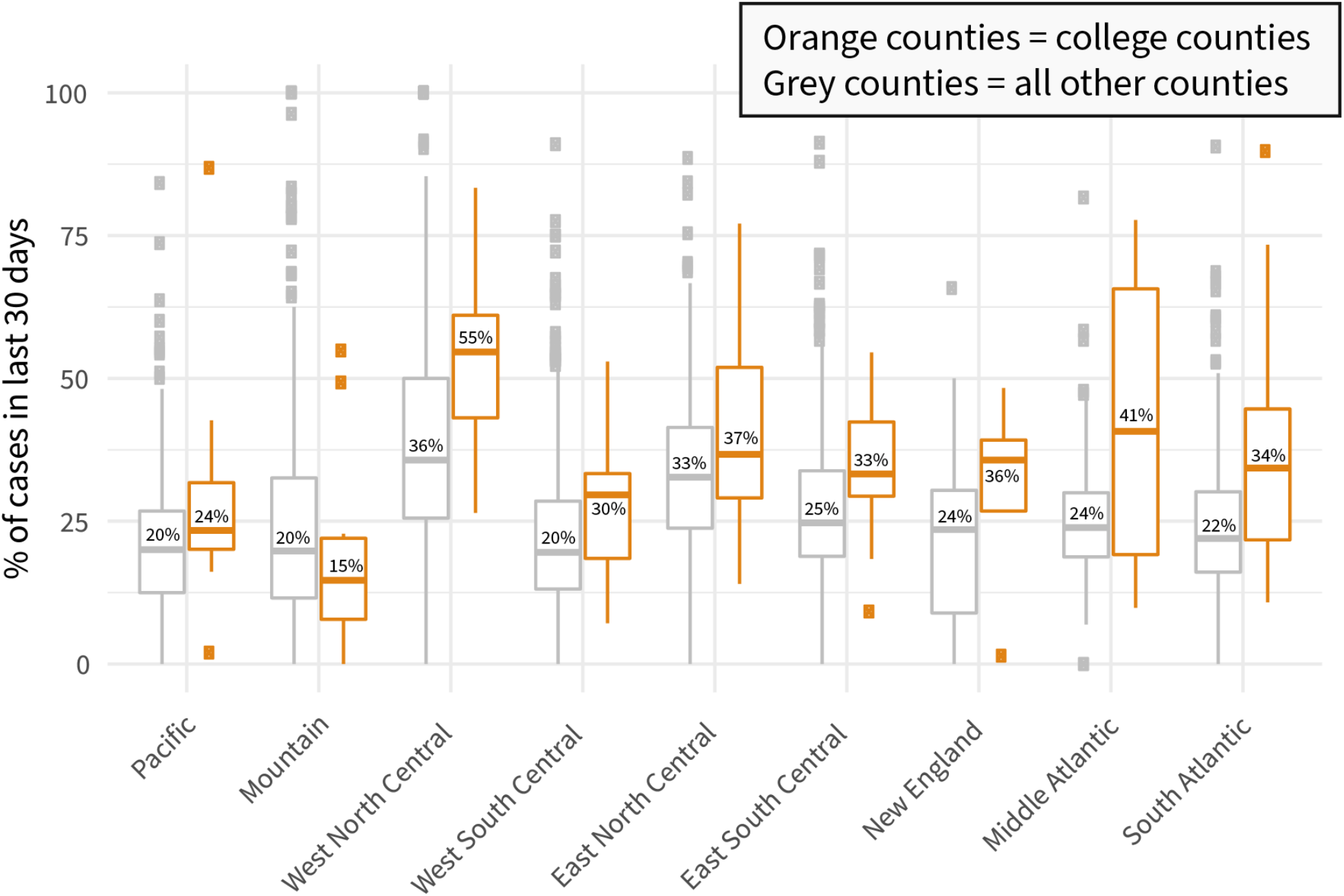
Box Plot of Difference in Median Return to Campus Case Rate Change by Census Division.

### Study Results

Descriptively, the new case rate varies notably within most census divisions, as shown in Exhibits 1, 2, and 3. In all but the Mountain Division, college counties have median rates higher than other counties. As Exhibit 2 shows, those differences range from 3.4% in the Pacific Division to 18.9% in the West North Central Division. The median new case rate is higher than 25% for college counties in all divisions except the Pacific and Mountain Divisions, indicating a surge in recent cases, with a maximum value of 54.6% in the West North Central Division. This may be partly due to a high new case rate for non-college counties (36%). Yet as the boxplots illustrate, this is only slightly higher than the neighboring East North Central Division (33%). In that division, new case rates for college counties are only 37%, 18% lower than in the West North Central Division.

**Exhibit 3:**
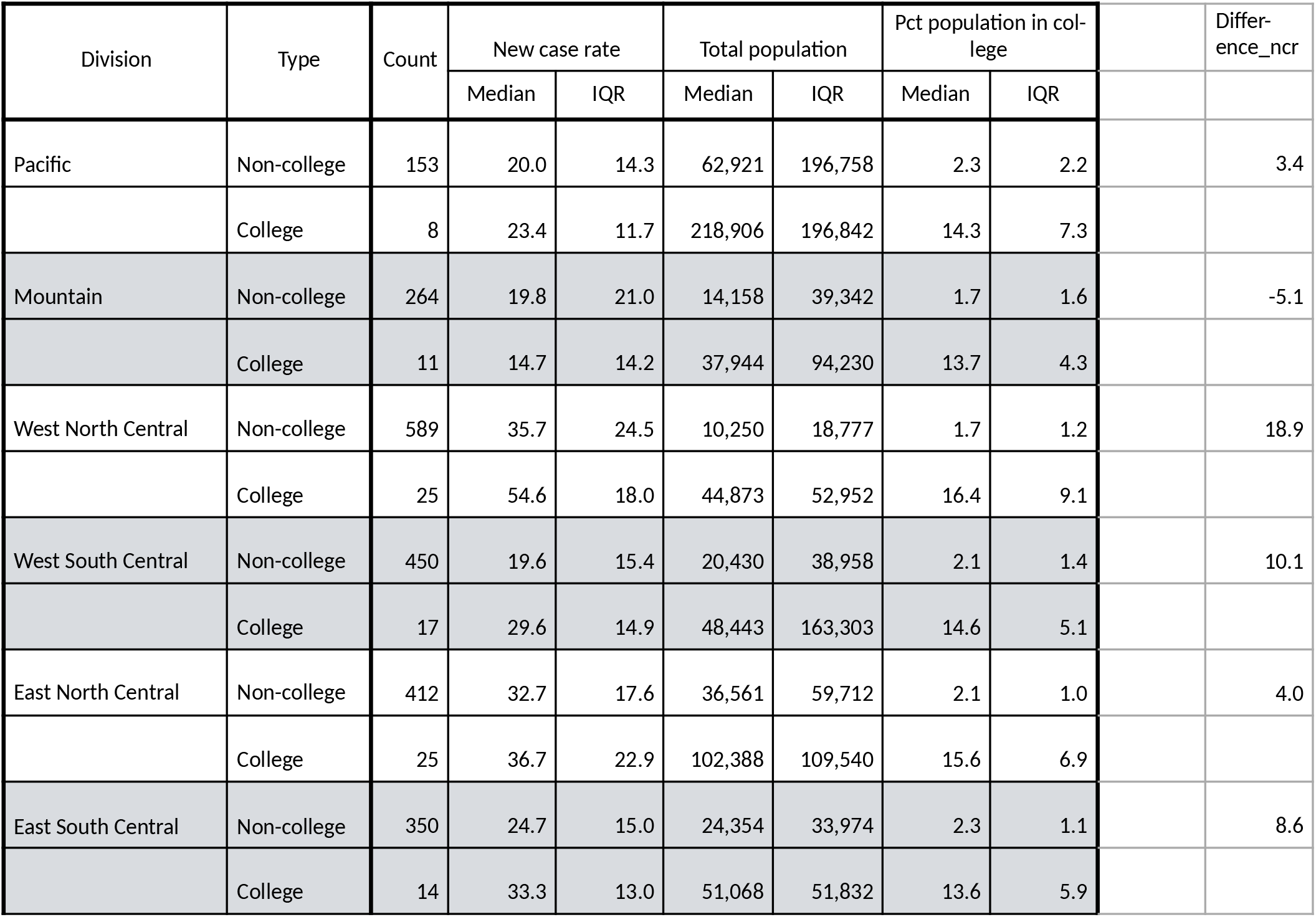
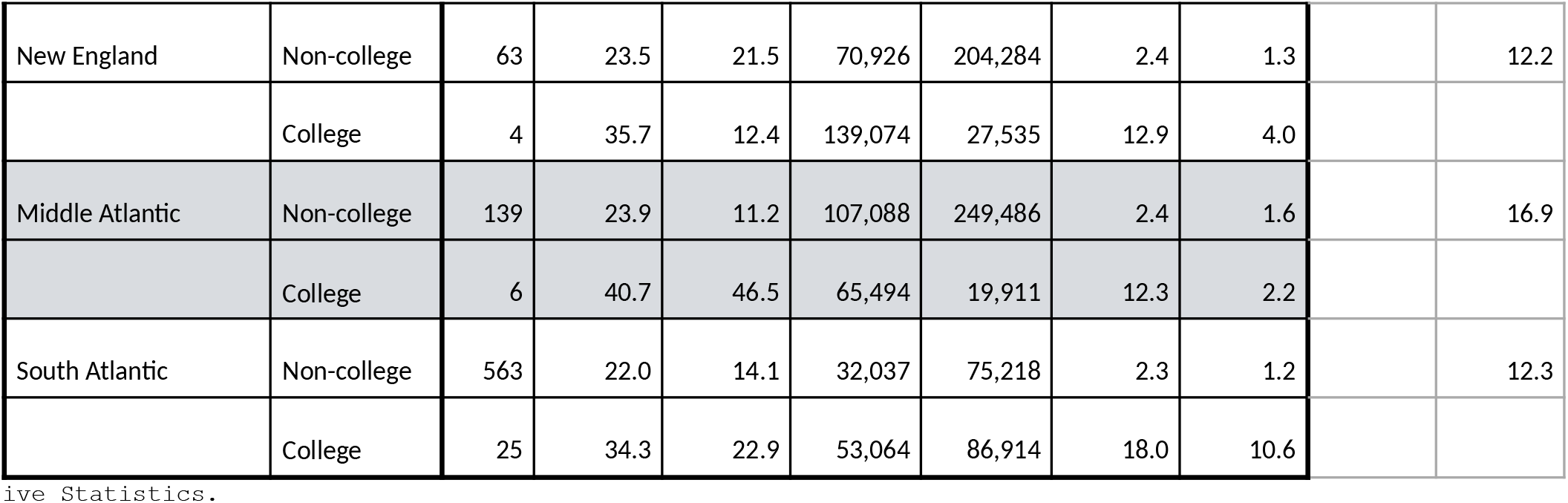
Table of Descriptive Statistics.

Exhibit 1 visualizes this through a map of county rates, with college counties highlighted as circles. The dark colors for many counties in the East and West North Central Divisions show the general surge of cases there, but almost all college counties have values at the high end of the classification range. In the East South Central and South Atlantic Divisions, new case rates are lower, but the college counties in these areas are still high relative to non-college counties.

The total population of college counties is consistently higher than non-college counties, with the only exception being the Middle Atlantic Division. This is largely due to the rural nature of most counties nationally. The median percentage of college students ranges from 1.7% to 2.4% in non-college counties. Lastly, college counties are most numerous in the West North Central and South Atlantic Divisions (25 in each) and least numerous in New England (4), though the rate in the latter (6% of 67 total counties) is higher than in the former (4% in each).

The results of our primary model (model 1, shown in Exhibit 4) indicate that college counties have a significant, positive association with higher new case rates, one that is 9.2% higher than other counties in their divisions, controlling for population size. This coefficient is slightly smaller (8.5%) in model 2, where the new case rate uses all reported cases as a baseline rather than the last 12 weeks.

**Exhibit 4:**
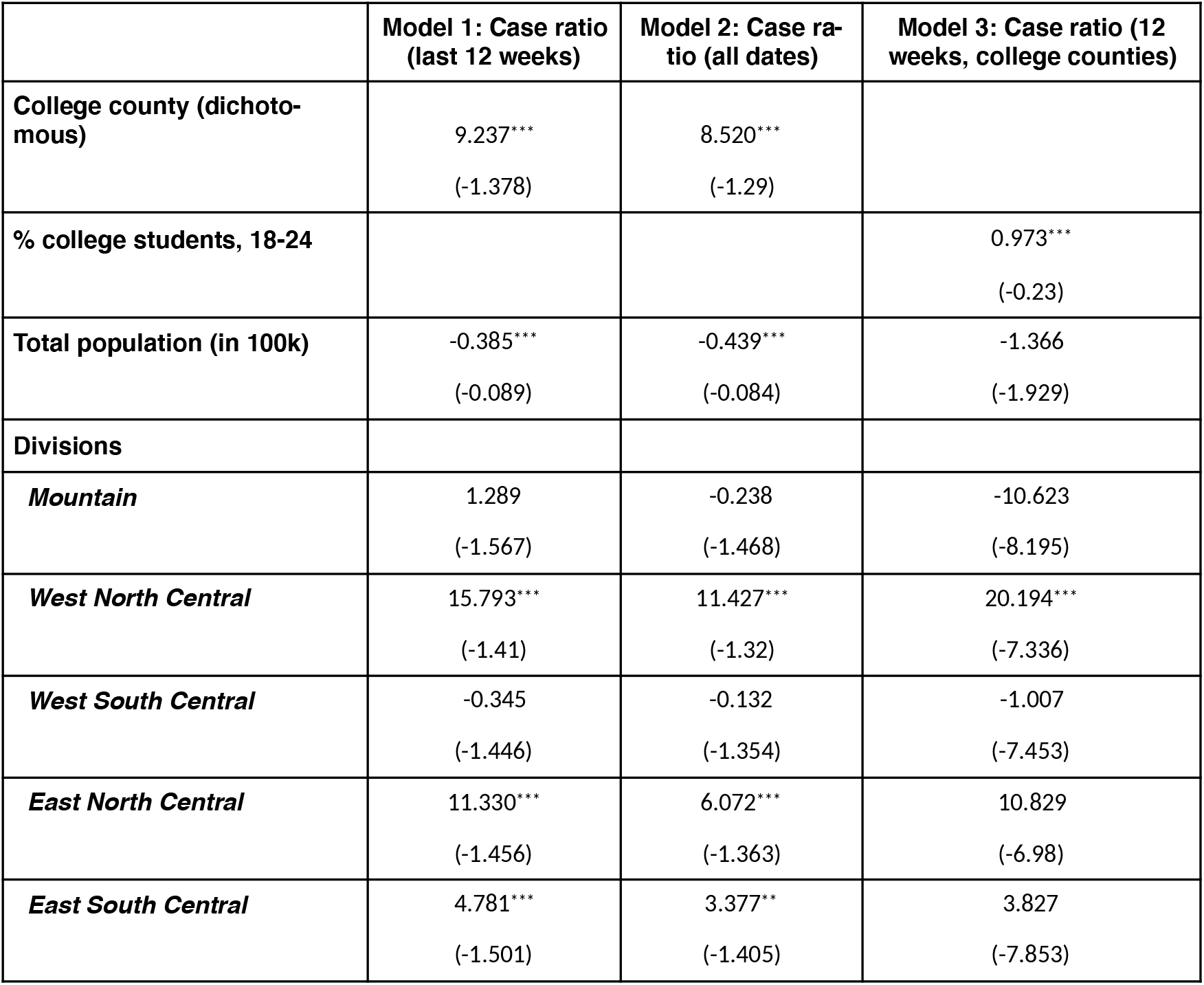
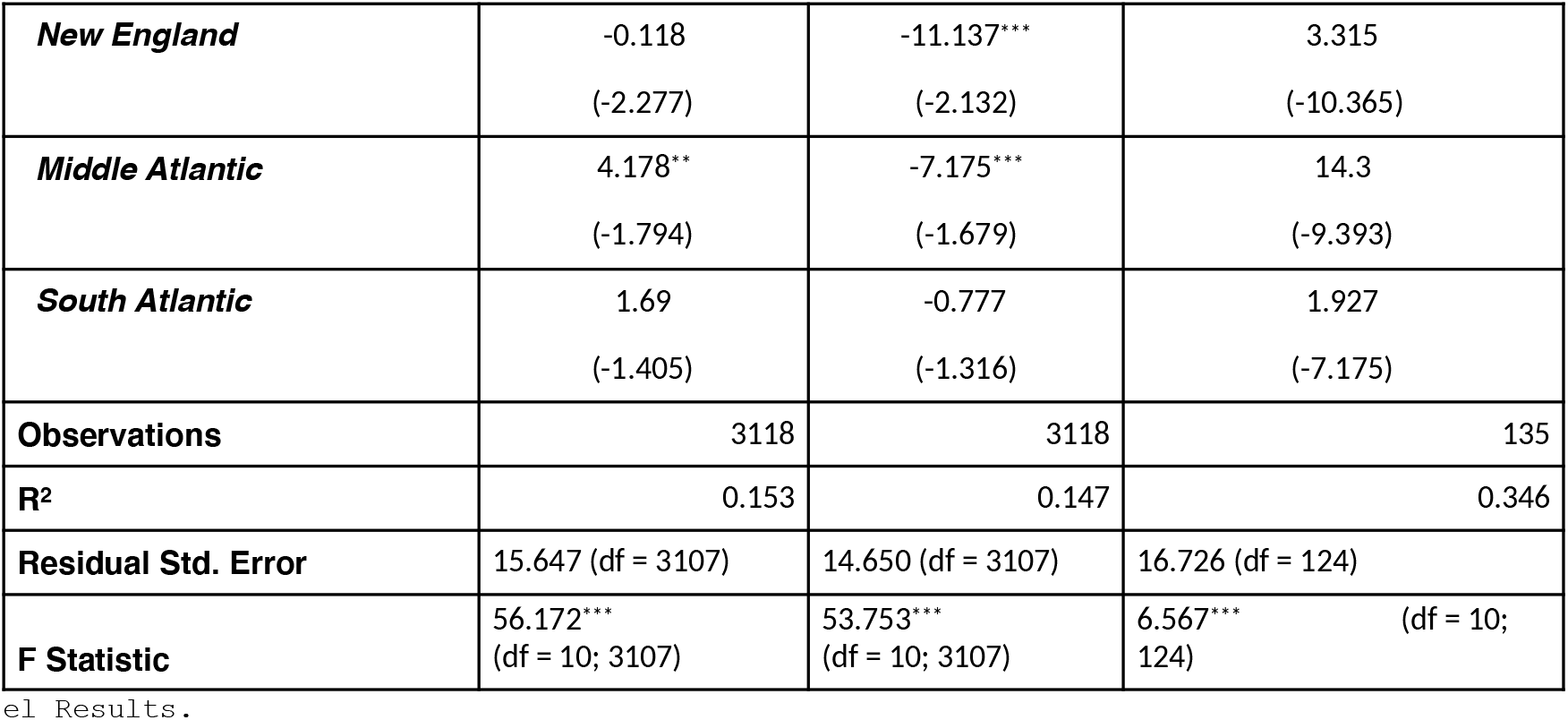
Model Results.

**Exhibit 5:**
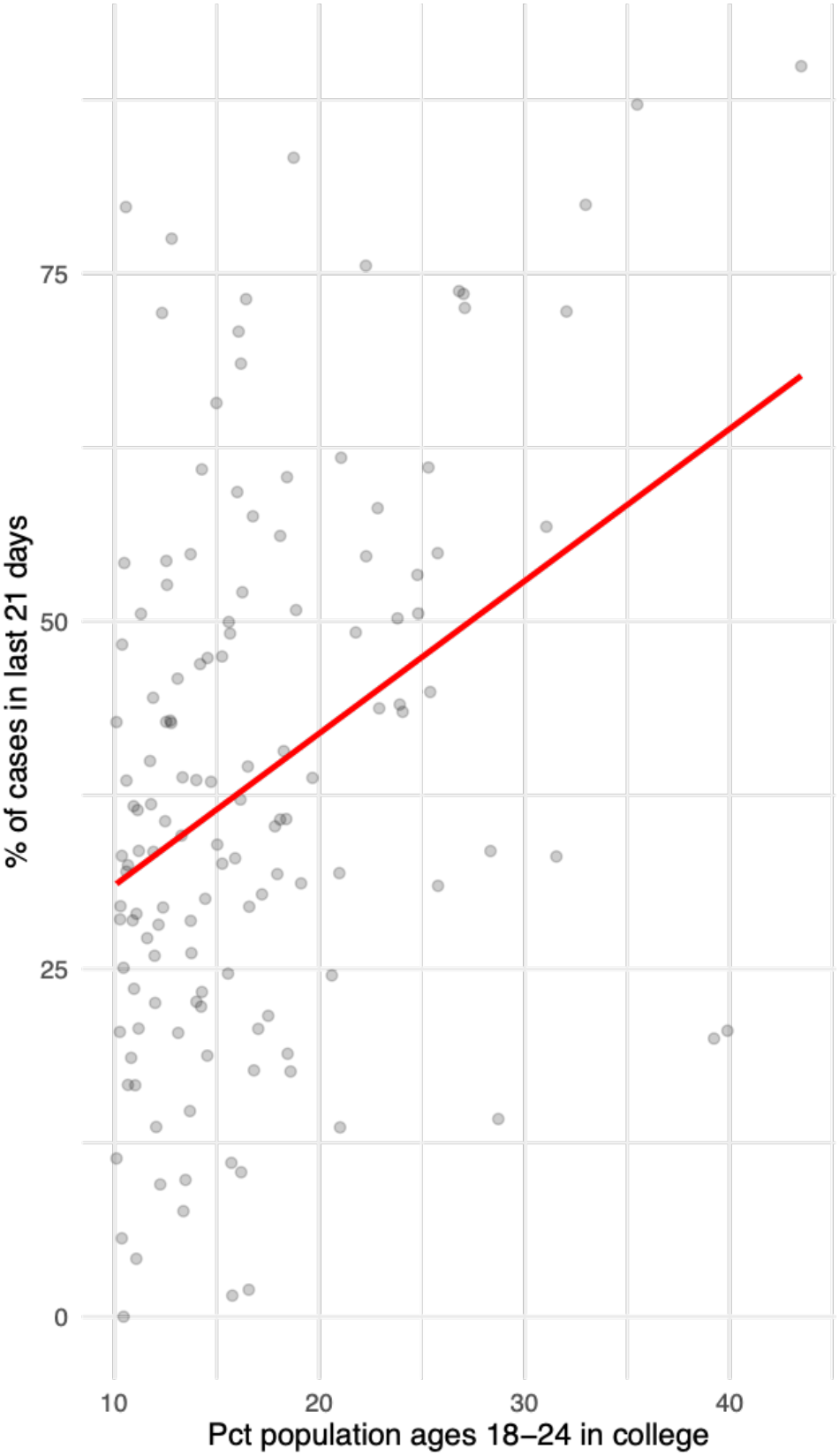
Graph of Model 3 Goodness of Fit.

The coefficient for total population is negative in both these models, which indicates that the new case rate is higher in counties with smaller population. The magnitude and direction of this coefficient is similar (−0.39, significant) even when the college county variable is removed. The control variables for division are significant in both models 1 and 2 in the West and East North Central, East South Central, and Middle Atlantic Divisions (the Pacific Division is used as a reference). These coefficients are mostly positive, as both Exhibits 1 and 2 show recent surges in the first three divisions. The R^2^ shows a modest fit (0.153) for these models.

When considering just college counties (model 3), model fit is much stronger (R^2^=0.346), as many counties with high rates but few college students have been removed. The coefficient for the rate of college students is positive and significant (0.97), indicating that new case rates increase approximately 1% for each additional 1% of college students in the population. Thus, case surges in college counties are greater in towns with a greater percentage of students.

## Discussion

We find support for the hypothesis that the return to college and university campuses across the United States is associated with significant increases in county-level new case rates, controlling for the county population and Census division. College counties (those with >10% of their population enrolled in colleges or universities) show significant increases in new case growth relative to counties not in this category. These findings serve as preliminary evidence of the effect of in-person return to college campuses on COVID19 viral transmission in the counties where these postsecondary institutions are located.

Our response to the pandemic as a society has rapidly evolved from full-scale lockdown in the first wave of infections, to a fully reopened economy during the summer that has remained open despite new waves of infections. In the beginning of the pandemic in the United States, many local and state jurisdictions adopted and implemented shelter-in-place (SIP) policies. These policies required that individuals stay at home unless conducting essential tasks (such as grocery shopping or picking up prescriptions) or performing essential work functions (such as healthcare provision).

During this time period, which spanned from mid-March to late April, over 94% of the American population was under a SIP order.^8^ The SIP policies and the weeks that followed their implementation meant that most individuals were socially isolated, with implications for individuals and households in experiencing increased loneliness and decreases in mental wellbeing.^9,10^ This also applied to students in the postsecondary education population. Colleges and universities shut down in-person operations and moved classes to virtual delivery.

After months of SIP policies, followed by a mixed landscape of less formal recommendations at state and local levels to maintain social distancing standards and to avoid large groups, colleges and universities have brought millions of students back to campuses across the country. The students could have a significant pent-up demand for social engagement and a desire for a return to normalcy, despite the persistence of the pandemic. Preliminary evidence of the effects of the pandemic and SIP policies show that “quarantine fatigue” was evident as early as mid-April.^11^ By the fall 2020 return to campus, quarantine and pandemic fatigue are likely much worse.

The success of limiting viral transmission rests on our ability and willingness as a society to comply with public health mitigation efforts, included but not limited to consistent mask wearing, social distancing, avoiding large groups (especially indoors), access to testing with timely results, compliance with contact tracing, and quarantine and isolation when exposed or testing positive. Policies within institutions/systems of higher education as well as college counties should attend to the specific developmental stage of young adulthood and leverage incentives for compliance accordingly.^12,13^ As many are now discovering, simply cajoling young people to stay home and not spend significant time with their peers is ineffectual and developmentally inappropriate.

### Limitations

Interpretation of our findings requires some caution. First, we would expect that part of the case increase in counties with institutions of higher education is simply a direct function of the population of those counties increasing with the return to campus. Second, there is significant variation in local and state-level policy response to the pandemic that is a potential source of omitted variable bias in our analysis. Third, post-secondary institutions are taking myriad approaches to their return to campus.

The work of Chris Marsicano and colleagues have documented the various approaches to this policy landscape. These range from full scale returns to campus with in-person instructions, to hybrid models combining the return to in-person combined with virtual learning, to completely moving instructions and university operations online for the duration of the 2020-2021 academic year.^14^ We are unable to account for the nuances of the variation in instructional model, in addition to the various approaches that are being taken to testing, contact tracing, and other pandemic mitigation strategies.

### Conclusion

Our analysis reveals a strong association between student return to campus and new case growth, specifically in “college counties” in the United States. For these counties, where at least 10% of the county population is comprised of college students, we see an average increase of 9% in the new case rate relative to non-college counties. These findings are consistent across the country at the U.S. Census Division level, with the Mountain Division as the only exception.

Return to campus case rates grow significantly in college counties when controlling for both the county population and the college or university population, and the findings are robust whether we estimate the models across the total pandemic observation period and the 60 days prior to campus return. These findings suggest that despite the efforts of colleges and universities to prepare for a safe return to campus, virus transmission has persisted among college students and is rapidly evolving in ways that has already created new “hot spots.” The science on COVID19 transmission is still developing, but thus far it has been established that the spread is a function of respiratory droplets from infected persons coming into contact with non-infected persons.

Given this reality, colleges and universities and the communities in which they are embedded must act quickly to intervene to slow the transmission before seasonal change and colder weather brings flu season. While it is expensive for postsecondary institutions to pivot and forgone revenue will come with its own host of consequences for both these institutions and the counties where they are located, COVID-19 transmission has already reached levels that could lead to overwhelm the healthcare systems of these college counties. Prioritizing public health demands that until effective vaccines or other anti-viral therapeutics are developed, policy approaches of postsecondary institutions and local county governments will have to become more collaborative and aggressive to control viral transmission.

## Data Availability

Data are available from the GitHub link posted below.

https://github.com/nytimes/covid-19-data/tree/master/colleges

## References

1. Courtemanche, C., Garuccio, J., Le, A., Pinkston, J., & Yelowitz, A. (2020). Strong Social Distancing Measures In The United States Reduced The COVID-19 Growth Rate: Study evaluates the impact of social distancing measures on the growth rate of confirmed COVID-19 cases across the United States. Health Affairs, 10–1377.

2. Roberts, Siobhan. (2020, March 27). How to Flatten the Curve on Coronavirus. New York Times. Accessed September 14, 2020 at: https://www.nytimes.com/article/flatten-curve-coronavirus.html.

3. College Crisis Initiative. Accessed September 7, 2020 at: https://collegecrisis.shinyapps.io/dashboard/.

4. Klompas, M., Baker, M. A., & Rhee, C. (2020). Airborne transmission of SARS-CoV-2: theoretical considerations and available evidence. Jama.

5. Wu, D., Wu, T., Liu, Q., & Yang, Z. (2020). The SARS-CoV-2 outbreak: what we know. International Journal of Infectious Diseases.

6. U.S. Census Bureau. American Community Survey Data. Accessed September 7, 2020 at: https://www.census.gov/acs/www/data/data-tables-and-tools/data-profiles/.

7. New York Times. Coronavirus (Covid-19) Data in the United States. 2020. Available from: https://github.com/nytimes/covid-19-data.

8. Dave, D., Friedson, A. I., Matsuzawa, K., & Sabia, J. J. (2020). When Do Shelter-in-Place Orders Fight COVID-19 Best? Policy Heterogeneity Across States and Adoption Time. Economic Inquiry.

9. Killgore, W. D., Cloonen, S. A., Taylor, E. C., & Dailey, N. S. (2020). Loneliness: A signature mental health concern in the era of COVID-19. Psychiatry Research, 113117.

10. Hsing, A., Zhang, J. S., Peng, K., Lin, W. K., Wu, Y. H., Hsing, J. C., … & Lounsbury, D. W. (2020). A Rapid Assessment of Psychological Distress and Well-Being: Impact of the COVID-19 Pandemic and Shelter-in-Place. Available at SSRN 3578809.

11. Zhao, J., Lee, M., Ghader, S., Younes, H., Darzi, A., Xiong, C., & Zhang, L. (2020). Quarantine Fatigue: first-ever decrease in social distancing measures after the COVID-19 pandemic outbreak before reopening United States. arXiv preprint 2006.03716.

12. Steinberg, L. (2007). Risk Taking in Adolescence: New Perspectives From Brain and Behavioral Science. Current Directions in Psychological Science, 16(2), 55–59. https://doi.org/10.1111/j.1467-8721.2007.00475.x

13. Albert, D., Chein, J., & Steinberg, L. (2013). The Teenage Brain: Peer Influences on Adolescent Decision Making. Current Directions in Psychological Science, 22(2), 114–120. https://doi.org/10.1177/0963721412471347

14. Marsicano, C., Felten, K., Toledo, L., & Buitendorp, M. (2020). Tracking Campus Responses to the COVID-19 Pandemic. APSA Preprints. doi:10.33774/apsa-2020-3wvrl

